# Convalescent Plasma in COVID-19. Mortality-Safety First Results of the Prospective Multicenter FALP 001-2020 Trial

**DOI:** 10.1101/2020.11.30.20218560

**Authors:** Raimundo Gazitúa, José Luis Briones, Carolina Selman, Franz Villarroel-Espíndola, Adam Aguirre, Roxana González-Steigmaier, Karina Cereceda, Mauricio Mahave, Betzabé Rubio, Pedro Ferrer-Rosende, Jorge Sapunar, Hugo Marsiglia, Ricardo Morales, Fernanda Yarad, María Elvira Balcells, Luis Rojas, Bruno Nervi, Jyh Kae Nien, Javier Garate, Carolina Prieto, Sofía Palma, Carolina Escobar, Josefina Bascuñan, Rodrigo Muñoz, Mónica Pinto, Daniela Cardemil, Marcelo Navarrete, Soledad Reyes, Victoria Espinosa, Nicolás Yáñez, Christian Caglevic

**Author notes:** **Corresponding author:** Raimundo Gazitúa, Instituto Oncológico Fundación Arturo López Pérez, Avenida Rancagua 795, 3rd Floor, Hematology Department, Santiago de Chile.

## Abstract

**Background:** The use of convalescent plasma (CP) to treat COVID-19 has shown promising results; however, its effectiveness remains uncertain. The purpose of this study was to determine the safety and mortality of CP among patients hospitalized with COVID-19.

**Study Design and Methods:** This multicenter, open-label, uncontrolled clinical trial is currently being conducted at nine hospitals in Chile. Patients hospitalized due to COVID-19 who were still within 14 days since symptom onset were classified into four groups: Patients with cancer and severe COVID-19. Patients with cancer and non-severe COVID-19. Patients with severe COVID-19 and patients with non-severe COVID-19 only. The intervention involved two 200-cc. CP transfusions with anti-SARS-CoV-2 IgG titers ≥ 1:320 collected from COVID-19-recovered donors.

**Results:** 192 patients hospitalized for COVID-19 received CP transfusions. At the first transfusion, 90.6% fulfilled the criteria for severity, and 41.1% required mechanical ventilation. 11.5% of the patients had cancer. Overall 7-day and 30-day mortality since the first CP transfusion was 5.7% and 16.1% respectively. There were no differences at either time point in mortality between the four groups. Patients on mechanical ventilation when receiving CP had higher mortality rates than those who were not (22.8% vs. 11.5%; p = 0.037). Overall 30-day mortality was higher in patients over 65 than in younger patients (p = 0.019). Severe adverse events were reported in four patients (2.1%) with an overall transfusion-related lung injury rate of 1.56%. No CP-related deaths occurred.

**Discussion:** CP is safe when used in patients with COVID-19 even when also presenting severity criteria or risk factors. Our mortality rate is comparable to reports from larger studies. Controlled clinical trials are required to determine efficacy.

**Registration:** NCT04384588

## Introduction

Severe acute respiratory syndrome coronavirus 2 (SARS-CoV-2) was first described in December 2019 in Wuhan, China and quickly spread across the globe, being declared a pandemic by the World Health Organization (WHO) on March 11, 2020 (**1**). The first case in South America was reported on February 26 in Brazil (**2**), while in Chile, the index case was confirmed on March 3, 2020 (**3**). Currently, 29% of the cases worldwide are concentrated in Latin America, one of the most affected areas in the world (**2**), which constitutes a serious health problem considering the complex socioeconomic and health characteristics of this region.

At the moment, there is neither a completely effective standard of care nor a proven effective vaccine against COVID-19. Many therapies have been evaluated against SARS-CoV-2 with mixed results. The combination of hydroxychloroquine and azithromycin in an early case series seemed promising (**4**), but randomized studies were unable to confirm its efficacy, and had an even higher incidence of adverse effects (**5-7**). The antiviral drug remdesivir provides clinical improvement yet no decrease in mortality (**8,9**). Moreover, antiviral drugs lopinavir/ritonavir were also shown to be ineffective against SARS CoV-2 (**10**). Only low doses of dexamethasone decrease the mortality rate in patients with mechanical ventilation and oxygen supplementation requirements (**11**).

Passive immunotherapy with convalescent plasma (CP) from individuals who have already recovered from infection has been safe and effective against previous viral outbreaks including the SARS-CoV-1, MERS-CoV, influenza A (H1N1), and the Ebola virus epidemics (**12-17**). The initial reports published on using CP to treat COVID-19 suggest that it confers clinical improvement, decreases viral load, and shortens mechanical ventilation duration (**18,19**). The use of CP is safe, with an incidence of related serious adverse events of less than 1% of treated patients, similar to the usual practice of transfusion medicine (**20**).

In this work, we report on the safety and in-hospital mortality of patients with COVID-19 treated with CP. Because patients with cancer commonly have impaired humoral immunity **(21)** we included a separate cohort of patients with cancer as part of our compassionate access protocol.

The study is a part of the NCT04384588 protocol.

## Patients and Methods

This is a multicenter, open-label, uncontrolled clinical trial. For data analysis purposes, the cohort was divided into four arms: A) Patients with cancer who meet the severity criteria; B) Patients with cancer who do not meet the severity criteria but have at least one poor prognostic factor; C) Patients without cancer who meet the WHO severity criteria; and D) Patients without cancer without severity criteria with at least two risk factors for poor prognosis (S1 Appendixes 1a, 1b, and 1c) (**20, 22, 23, 24**). The criteria used to define patients with cancer are listed in S2 Appendix.

A case of COVID-19 was defined as a patient with clinically compatible symptoms (**25,26**) (S3 Appendix) and an RT-qPCR-based confirmation of SARS-CoV-2 from a nasopharyngeal swab or with a chest CT scan compatible with COVID-19 (**27**).

This project was developed and executed in accordance with Chilean Law No. 20,120 (“On scientific research in the human being, its genome, and prohibitis human clonation” : All scientific research on a human being must have his prior, express, free and informed consent, or, failing that, that of the person who must supply his will in accordance with the law.”); Law No. 20,584 (“Regulates the rights and duties that people have in relation to actions related to their health care”); Law No. 19.628 (“On protection of prívate life”); CIOMS Guidelines, Declaration of Helsinki, Nuremberg code, The Universal Declaration of Human Rights and The International Covenant on Civil and Political Rights

This protocol was approved by our local ethics committee: Comité Ético Científico Fundación Arturo López Pérez, on April 7^th^, 2020. Informed consent was obtained from all patients or a representative when the patients were unable to sign themselves due to their health condition. A total of 41 patients from the NCT04375098 trial were included in this report through a collaborative network.

### Study population

Patients were enrolled between May 1 and August 7, 2020 in nine hospitals from four Chilean cities: Instituto Oncológico Fundación Arturo López Pérez (FALP) (Santiago, Metropolitan Region), Pontificia Universidad Católica de Chile (Santiago, Metropolitan Region), Clínica Dávila (Santiago, Metropolitan Region), RedSalud (Santiago, Metropolitan Region), Hospital del Trabajador (Santiago, Metropolitan Region), Hospital Dipreca (Santiago, Metropolitan Region), Hospital Clínico de Magallanes (Punta Arenas, Magellan Region), Clínica Alemana Temuco (Temuco, Araucania region), Hospital Regional de Talca (Talca, Maule Region).

### Participation criteria

Patients were eligible to receive CP if they were at least 18 years old with a confirmed diagnosis of severe SARS-CoV-2 infection or non-severe COVID-19 with two or more poor prognostic factors within the first 14 days from symptom onset. Patients with a known allergy to previous plasma infusions, multiple organ dysfunction, active intracerebral hemorrhage, disseminated intravascular coagulation requiring transfusion, acute respiratory distress syndrome, or with active cancer and a survival expectancy less than 1 year (S4 Appendix) were excluded.

### Plasma collection, processing, and conservation

Plasma was obtained from voluntary donors registered through a website (https://www.donantecovid.cl/) who were diagnosed with COVID-19 by RT-PCR from a nasopharyngeal swab sample or who presented clinically compatible symptoms and tomographic findings. Before the donation, patients had to be asymptomatic for at least 21 days with two negative RT-qPCR tests on two consecutive days or asymptomatic for at least 28 days. All donors underwent a blood-bank selection survey plus microbiological screening for HIV, hepatitis-B and -C, syphilis, HTLV-I and -II, and Chagas. In addition, a nucleic acid amplification test (NAAT) was performed for hepatitis-B, -C, and HIV. All women, with or without a history of pregnancy, were tested for anti-HLA antibodies. Anti-SARS-CoV2 antibodies were measured, and only donors with IgG ≥ 1:320 (ELISA Euroimmun®) were selected. Plasma was collected by apheresis (Trima Accel ® or Spectra Optia, Terumo®) and the volumes obtained were pooled and divided into doses of 200 mL each. CP units were stored at −40°C.

### Convalescent plasma transfusion

Each patient received two 200-mL units 24 hours apart. Units were selected according to blood type and Rh compatibility. The transfusion procedure took 1 to 2 hours, according to the treating physician’s preference. Premedication once administrated with clorpheniramine i.v. 10mg and acetaminophen 1 gr. was also permitted.

### Variables analyzed

This report analyzed the primary outcomes of the FALP project COVID 001-2020 (NCT04384588): In-Hospital Mortality and Adverse Event Occurrence.

### Statistical analysis

Demographic and epidemiological variables are described in frequency and percentage, while continuous variables by their median and interquartile range. The relationships between the categorical variables were evaluated using the Chi-square test. When the expected frequency for a combination of variables was less than 5, Fischer’s exact test was perfomed. Overall survival was defined as the time from the first CP transfusion to the time of death or date of the last follow-up. Survival probability was estimated using the Kaplan-Meier method and the difference in survival between the groups was examined using the log-rank test. In addition, the Cox proportional-hazards model was applied to determine possible risk factors associated with mortality. A crude hazard ratio (HR) and an HR adjusted for sex, age, ventilation, days of hospitalization, and days of symptom duration until transfusion were estimated along with their 95% confidence intervals (Cis). A p-value under 0.05 was considered statistically significant. Statistical analyses were performed with STATA and R program v 3.6.0. (RCore Team, 2020, Vienna, Austria).

## Results

### Patient characteristics

Between May 1 and August 7, 2020, 192 patients from nine hospitals in four different regions of Chile enrolled in this study (Figure 1). Table 1 details the clinical and demographic characteristics of these patients. The median follow-up was 62 days (IQR: 34.8 to 88.0) while the median age was 59 years old (IQR: 49.8 to 67.0); 90.6% of the population met the severity criteria, 71.9% were hospitalized in an Intensive Care Unit, and 41.1% received mechanical ventilatory support, which was invasive in 30.7% of cases. The median SOFA score was 3 (IQR: 2 to 5) while the median PaO2/FiO2 was 181 (IQR: 127 to 273). The most common comorbidities were hypertension (59.0%) and diabetes mellitus (56.0%). Cancer was present in 11.5% of the cohort (n = 22)

**Table 1.** Baseline demographics and clinical characteristics.

|  |  |
| --- | --- |
| Age |  |
| Age – yr (median, IQR) | 59.0 (49.8 - 67.0) |
| Sex – no. (%) |  |
| Male | 135 (70.3) |
| Female | 57 (29.7) |
| Ethnic group – no. (%) |  |
| Hispanic or Latino | 180 (93.8) |
| Caucasian | 12 (6.2) |
| Severity – no. (%) |  |
| Severe | 174 (90.6) |
| Non-severe with RF | 18 (9.4) |
| O<sub>2</sub> requirement – no. (%) |  |
| No oxygen | 20 (10.4) |
| Oxygen | 172 (89.6) |
| Ventilatory support – no. (%) |  |
| Mechanic ventilation | 79 (41.1) |
| No mechanic ventilation | 113 (58.9) |
| Ventilatory support type – no. (%) |  |
| Invasive ventilation (IV) | 59 (30.7) |
| Non-invasive ventilation (NIV) | 20 (10.4) |
| O<sub>2</sub> support device – no. (%) |  |
| High-flow nasal cannula oxygenation | 35 (18.2) |
| Oxygen face mask | 11 (5.7) |
| Nasal oxygen cannula with prongs | 47 (24.5) |
| Hospitalization unit (at enrollment) – no. (%) |  |
| Critical care unit | 138 (71.9) |
| Basic room | 54 (28.1) |
| Comorbidities – no. (%) |  |
| Hypertension | 59 (30.7) |
| Diabetes | 56 (29.2) |
| Obesity | 41 (21.4) |
| Cancer | 22 (11.5) |
| EPOC | 10 (5.2) |
| Coronary cardiopathy | 5 (2.6) |
| HIV/AIDS | 0 (0) |
| Blood type – no. (%) |  |
| O | 113 (58.9) |
| A | 58 (30.2) |
| B | 19 (9.9) |
| AB | 2 (1.0) |
| SOFA |  |
| Sofa score at enrolment (median, IQR) | 3 (2-5) |
| PAFI |  |
| PaO<sub>2</sub>/ FiO<sub>2</sub> (median, IQR ) | 182 (127-273) |

**Figure 1.**
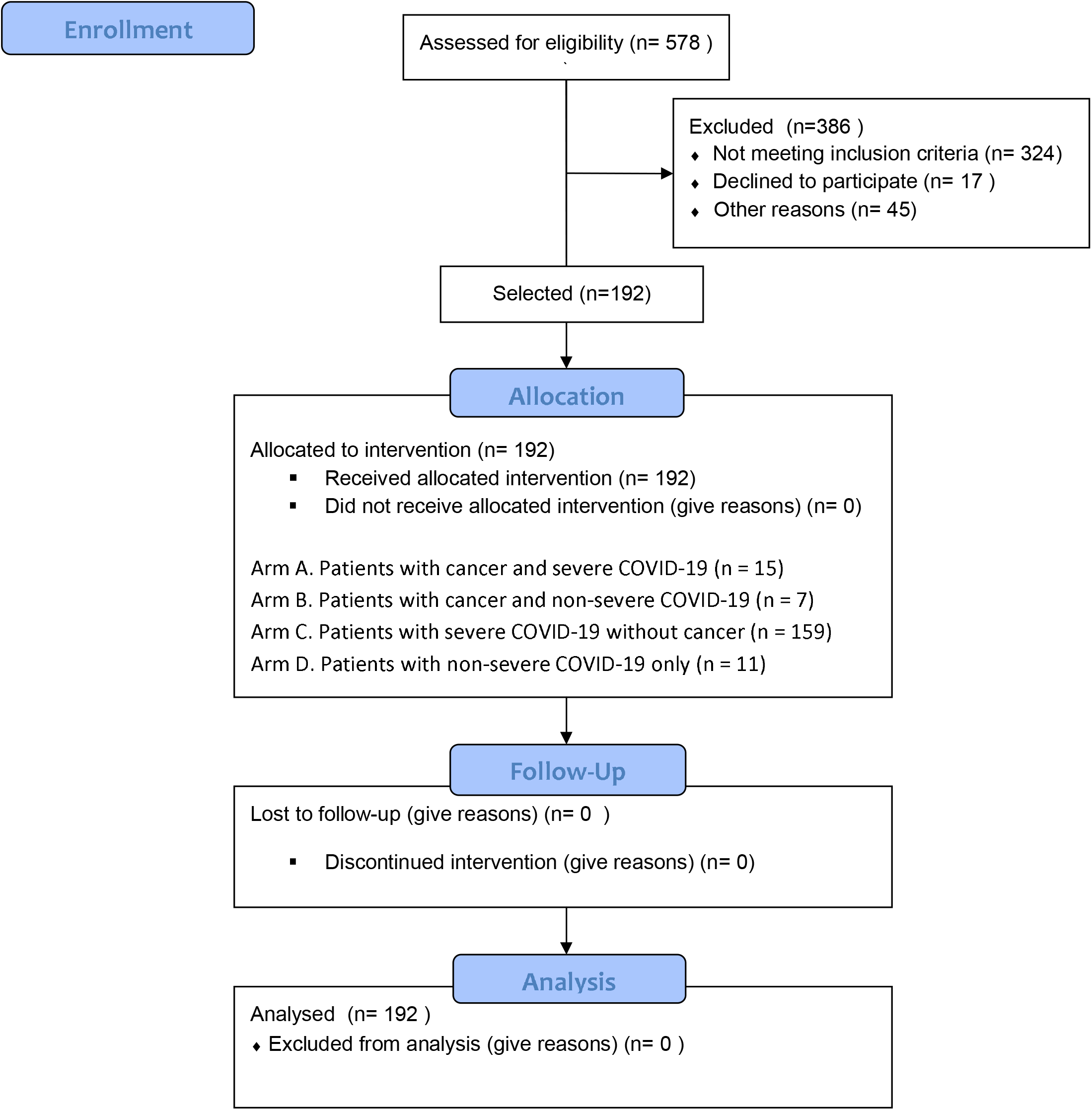
Study Flow Diagram.

### Outcomes

The mortality rates 7 and 30 days after CP administration were 5.7% and 16.1%, respectively (Figure 2). There were no significant differences in mortality related to the timing of the initial CP administration. Patients who received CP within the first 7 days of symptom onset had a 7-day mortality rate of 9.2% (95% CI, 4.1% to 17.3%) versus. 2.9% (95% CI, 0.6% to 8.1%; p = 0.06) for those who received CP between 8 and 14 days. Patients who received CP within the first 7 days of symptom onset had a 30-day mortality rate of 17.2% (95% CI, 10.0% to 26.8%) versus. 15.2% (95% CI, 9.0% to 23.6%; p = 0.707) for those who received CP between 8 and 14 days

**Figure 2.**
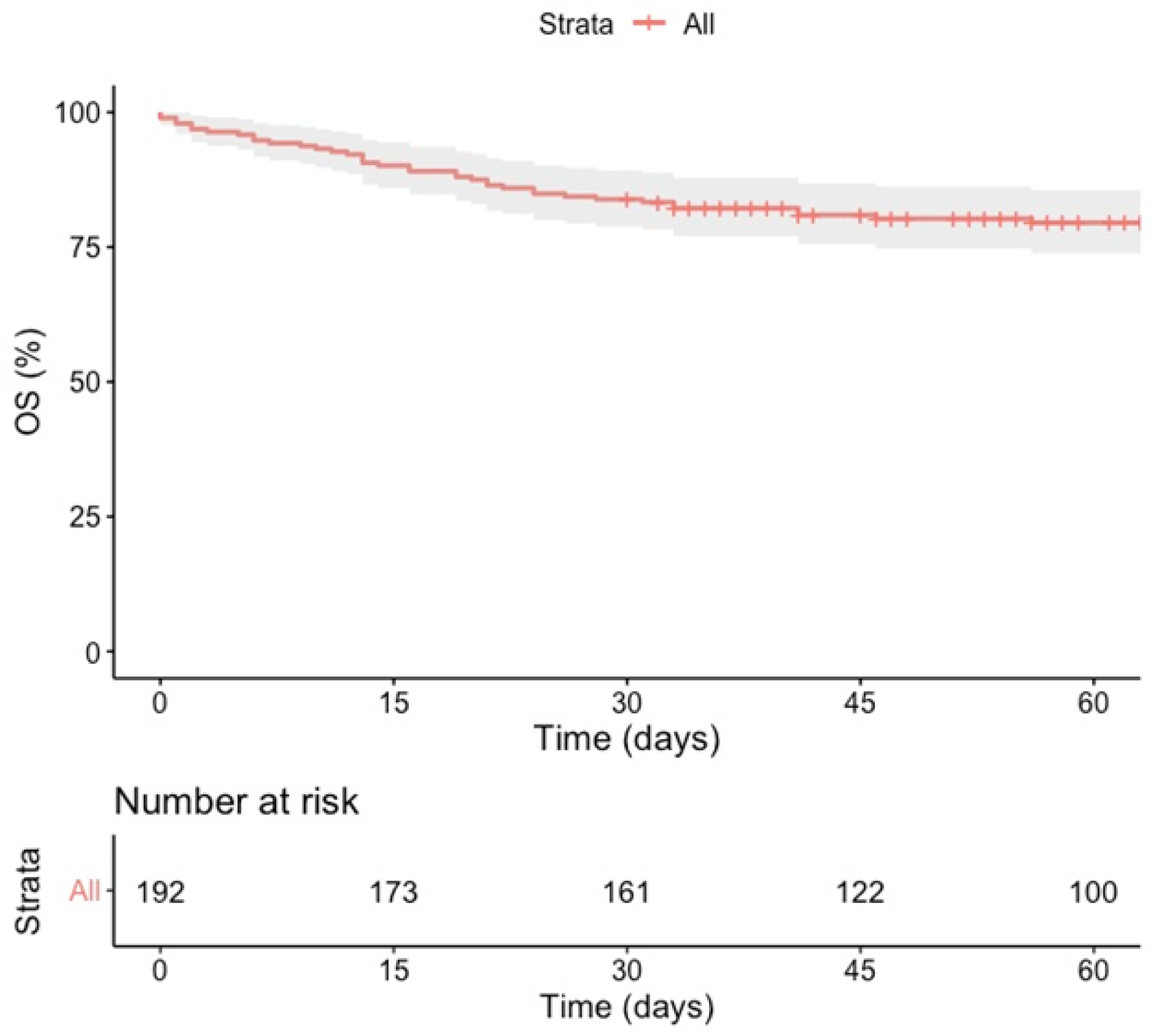
Time from enrollment to death. A Kaplan-Meier estimate of overall survival.

Patients who received plasma during the first 3 days of hospitalization did not register any significant differences in terms of 7- or 30-day mortality compared with those transfused after 3 days: 4.7% (95% CI, 1.7% to 9.8%) vs. 7.9% (95% CI, 2.6% to 17.6%; p = 0.358), and 14.0% (95% CI, 8.5% to 21.2%) vs. 20.6% (95% CI, 11.5% to 32.7%; p = 0.237) respectively. Patients receiving mechanical ventilatory support at the time of CP administration had a higher 30-day mortality compared with patients who did not require mechanical ventilation: 22.8% (95% CI, 14.1% to 33.6%) vs. 11.5% (95% CI, 6.3% to 18.9%; p = 0.037) (Table 2).

**Table 2.**
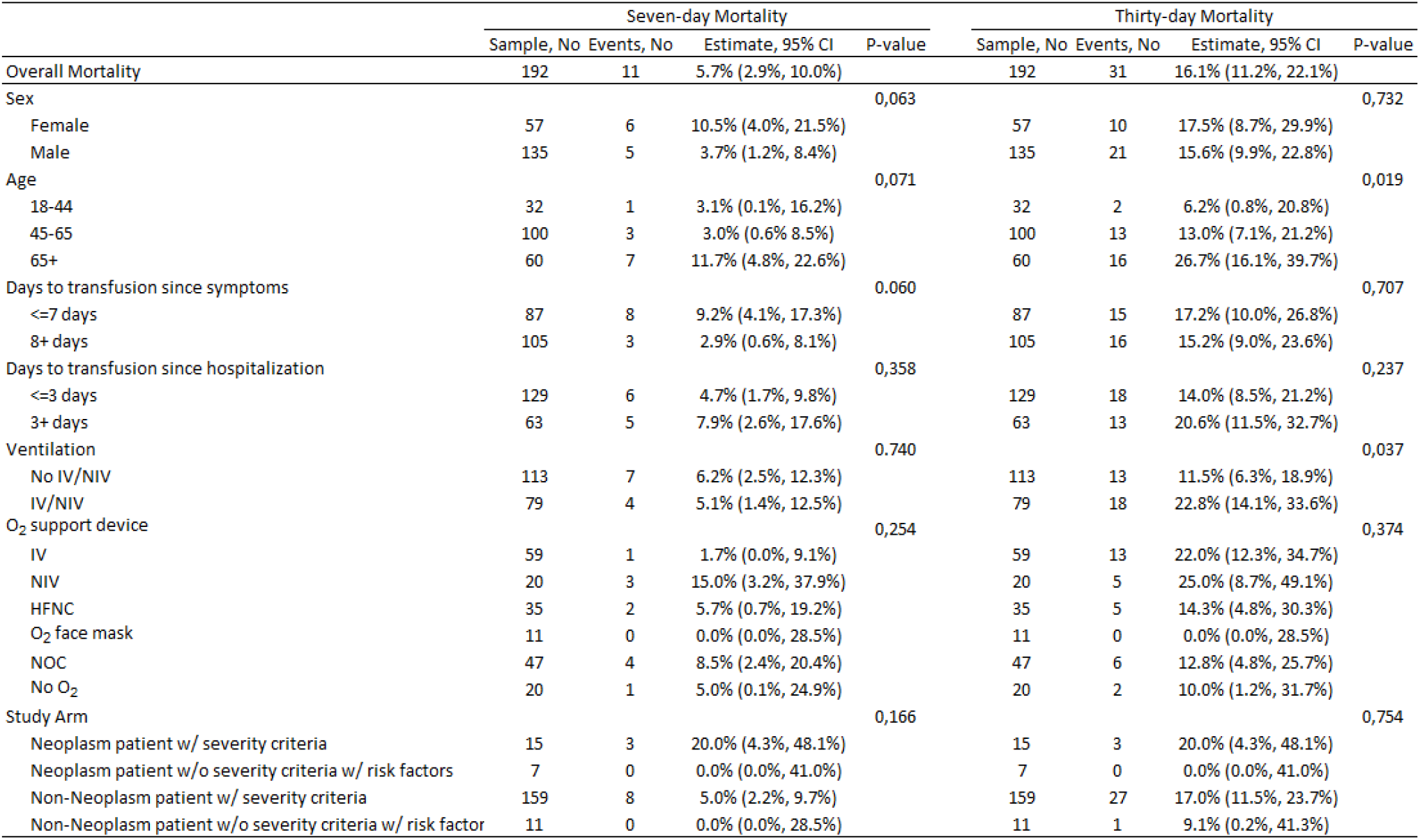
Seven and thirty-day mortality characteristics.

The trial arm with the highest mortality rate was the cancer and severe COVID group with 20.0% after 30 days (n = 3). The severe COVID only group had a 17.0% mortality rate. There were no deaths in the cancer and non-severe COVID group while the non-severe COVID only group registered one death (9,1%). There were no statistically significant differences between the different study groups in terms of 30-day mortality (Figure 3).

**Figure 3.**
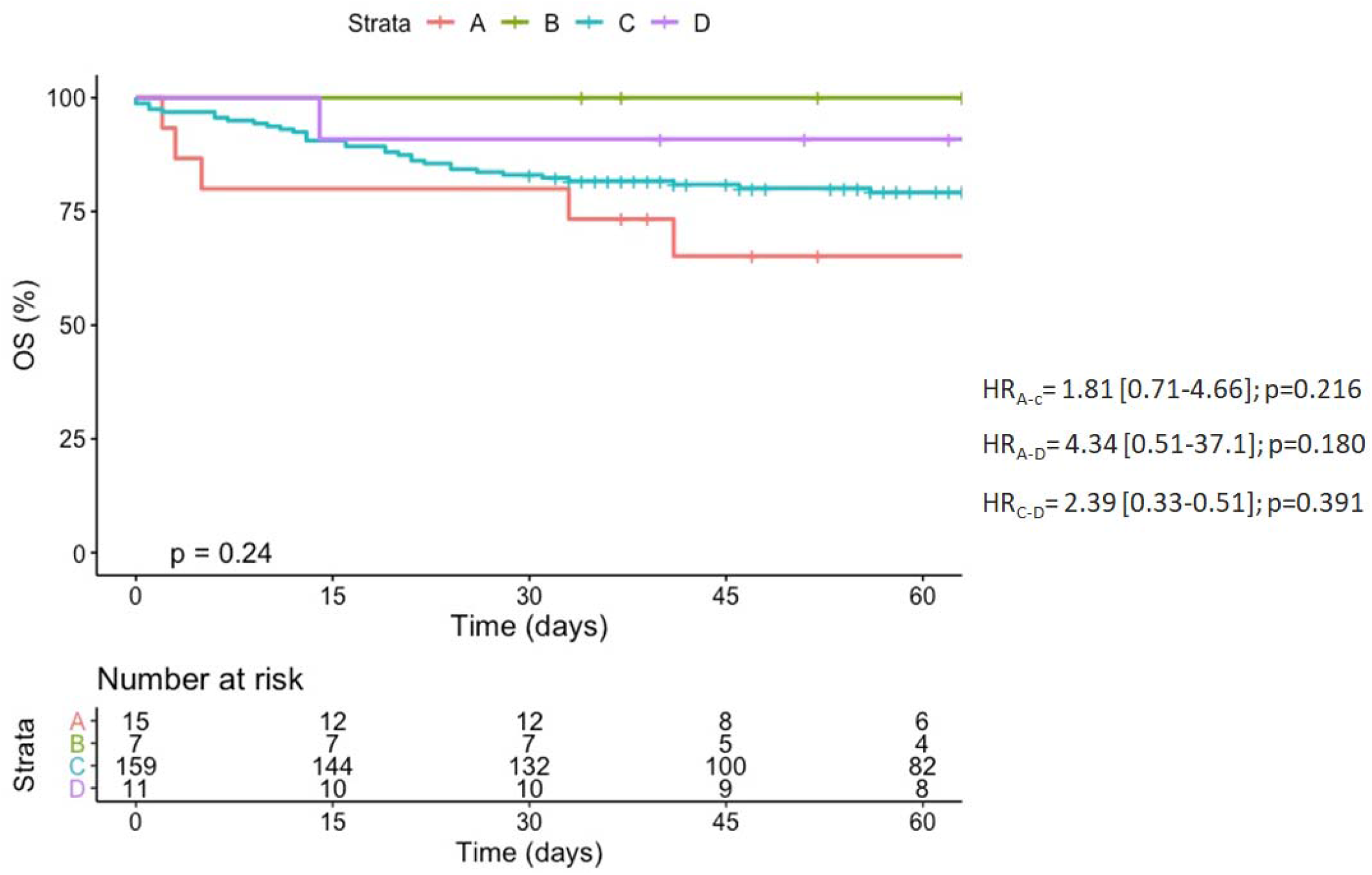
Overall Survival according to study arm. A Kaplan-Meier estimate of the overall survival between the study groups. (A) Patients with cancer and severe COVID-19. (B) Patients with cancer and non-severe COVID-19. (C) Patients with severe COVID-19 without cancer. (D) Patients with non-severe COVID-19 only.

Patients aged over 65 years old showed higher mortality (p = 0.019) compared with younger patients: 26.7% vs. 13.0% (45-65 years old) and 6.2% (18-44 years old) (Figure 4). There were no differences in mortality by sex (p = 0.326), ABO group (p = 0.89), or cancer status (p = 0.690). Age over 65 years old and mechanical ventilation at the time of CP administration were the two risk factors significantly associated with mortality (Table 3).

**Table 3.** Risk Factors Associated with Mortality Using a Cox Regression Model.

|  | Crude HR (95% CI) | P-value | Adjusted HR (95% CI) | P-value |
| --- | --- | --- | --- | --- |
| Sex |  |  |  |  |
| Female | 1 (Ref) |  | 1 (Ref) |  |
| Male | 0.78 (0.40-1.53) | 0,471 | 0.77 (0.38-1.57) | 0,471 |
| Age |  |  |  |  |
| 18-44 | 1 (Ref) |  | 1 (Ref) |  |
| 45-65 | 2.52 (0.58-11.0) | 0,221 | 2.10 (0.47-9.32) | 0,330 |
| 65+ | 6.67 (1.56-28.5) | 0,010 | 6.30 (1.45-27.32) | 0,014 |
| Days to transfusion since symptoms |  |  |  |  |
| <=7 days | 1 (Ref) |  | 1 (Ref) |  |
| 8+ days | 1.00 (0.53-1.89) | 0,991 | 0.73 (0.34-1.57) | 0,424 |
| Days to transfusion since hospitalization |  |  |  |  |
| <=3 days | 1 (Ref) |  | 1 (Ref) |  |
| 4+ days | 1.60 (0.84-3.04) | 0,155 | 1.31 (0.60-2.82) | 0,496 |
| Ventilation |  |  |  |  |
| No IV/NIV | 1 (Ref) |  | 1 (Ref) |  |
| IV/NIV | 2.30 (1.20-4.40) | 0,012 | 2.72 (1.24-5.54) | 0,006 |

**Figure 4.**
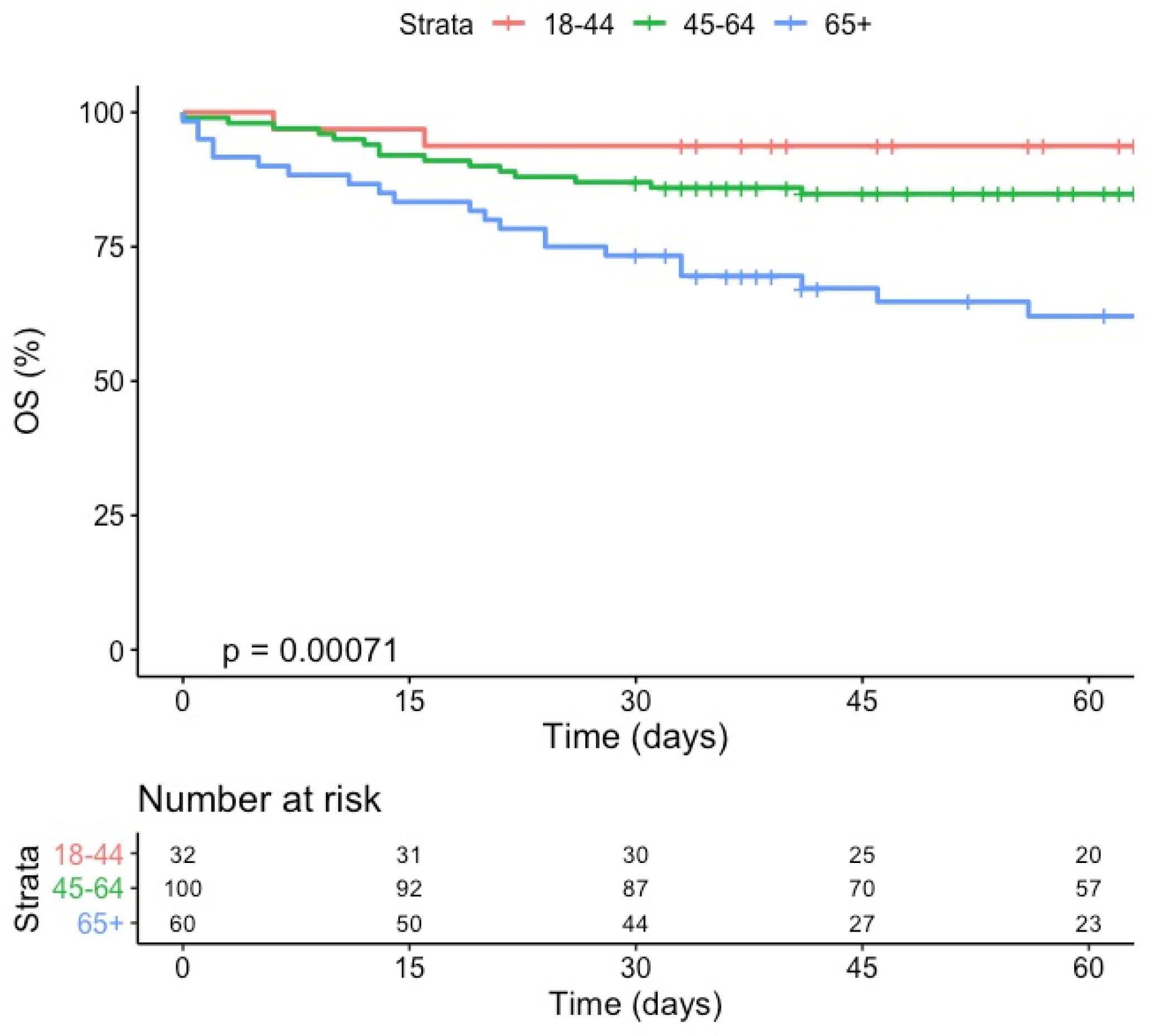
Overall survival by age. A Kaplan-Meier estimate of overall survival according to age group.

### Adverse events

We registered a total of 11 CP transfusion-related adverse events (2.9%), four (1.1%) of which were considered to be serious adverse events (SAEs) related to CP transfusion including three cases of transfusion-related acute lung injury (TRALI) and one case of thrombocytopenia. No CP transfusion-related deaths were reported (Table 4). The adverse events are summarized and described in Table 5.

**Table 4.** Adverse Events Related to the Use of Convalescent Plasma (CP)

| Adverse Event | Any Grade | Grade 3, 4 or 5* | SAE | Event/ Patient | Event / Transfusion |
| --- | --- | --- | --- | --- | --- |
| TRALI | 3 | 3 | 3 | 1.56 % (3/192) | 0.80 % (3/373) |
| Thrombocytopenia | 1 | 1 | 1 | 0.52 % ( 1/192) | 0.27% ( 1/373) |
| Fever | 5 | 0 | 0 | 2.60% ( 5/192) | 1.34% (5/373) |
| Skin - Rash | 2 | 0 | 0 | 1.04% (2/192) | 0.53% (2/373) |
| Total AE | 11 | 4 | 4 | 5.72% ( 11/192) | 2.94% ( 11/373) |
| Total SAE |  |  |  | 2.08% ( 4/192) | 1.07% (4/373) |
| Treatment related death |  |  |  | 0% ( 0/192) | 0% ( 0/373) |

\*CTCAE v5.0 ; SAE = Serious adverse event; TRALI = Transfusion related acute lung injury; AE = Adverse Events

**Table 5.** Adverse Events Registered During the Study.

| Adverse Events | Any Grade | Patients with AE | events per patient | AE Grade 3, 4 or 5 | Patients with AE Grade 3, 4 or 5 | events per patient |
| --- | --- | --- | --- | --- | --- | --- |
| System | n | n (%) | (range) | n | n (%) | (range) |
| Infectious | 62 | 43 (22.3) | (1-4) | 33 | 24 (12.5) | (1-3) |
| Respiratory | 20 | 18 (9.4) | (1-2) | 6 | 6 (3.1) | 1 |
| Renal | 17 | 14 (7.3) | (1-2) | 10 | 10 (5.2) | 1 |
| Cardiac | 17 | 16 (8.3) | (1-2) | 3 | 3 (1.6) | 1 |
| Gastrointestinal | 16 | 11 (5.7) | (1-2) | 1 | 1 (0.5) | 1 |
| Vascular-Thromboembolism | 11 | 11 (5.7) | 1 | 5 | 5 (2.6) | 1 |
| Neurologic | 8 | 8 (4.2) | 1 | 4 | 4 (2.1) | 1 |
| Fever | 5 | 5 (2.6) | 1 | 0 | 0 (0) | 1 |
| Skin | 4 | 4 (2.1) | 1 | 2 | 2 (1.0) | 1 |
| Hepatic | 3 | 3 (1.6) | 1 | 0 | 0 (0) | 1 |
| Hematologic | 2 | 2 (1.0) | 1 | 1 | 1 (0.5) | 1 |
| Reumatologic | 1 | 1 (0.5) | 1 | 0 | 0 (0) | 1 |
| Total | 166 | 136 (70.8) | (1-4) | 65 | 56 (29.2) | (1-3) |

AE = Adverse Event

## Discussion

To the best of our knowledge, this is the first report of in-hospital mortality and treatment safety from a Latin-American multicenter study on patients with COVID-19 who received CP. Although this study did not aim to demonstrate the efficacy of CP, our reported 7- and 30-days mortality rates are comparable with those reported by the Mayo Clinic in the largest CP study to date, which showed 7- and 30 day-mortality rates of 10.5% and 24.5%, respectively. Unlike the series published by Joyner et al. (2020), our cohort did not display differences in mortality associated with CP administration during the first 3 days of hospitalization compared to patients who were transfused later (**22**).

Mortality rates reported in patients with cancer and COVID-19 infection range from 30% to 39% (**28,29**). Many patients with cancer are immunocompromised as a consequence of their underlying disease and an associated treatment regimen that often includes myelosuppressive chemotherapy, immunosuppressive agents, and radiation, which may increase mortality (**30**). In our series, patients with cancer had a 30-day mortality incidence of 13.6%, similar to patients without cancer and lower than reported in other cohort studies of patients with cancer and COVID-19 infection. This low mortality rate may indicate that patients with an impaired humoral and cellular response may benefit from passive antibody administration impeding viral replication and modulating the inflammatory events associated with COVID-19. In contrast to our results, a recent study on the use of CP in patients with cancer reported a mortality rate of 41.7% (31). One important difference between the two cohorts is the proportion of patients with hematological malignancies; 58,1% was reported by Remblay et al, versus 36,3% from our trial. This particular subgroup seems to have higher mortality rates than patients with solid tumors, which is an interesting issue that warrants additional investigation.

As expected, elderly patients and patients requiring mechanical ventilation had significantly higher mortality rates in our study. These findings are consistent with previous evidence in patients with and without COVID-19 (**32,33**).

The main endpoint of our study was a safety assessment of the occurrence and type of CP-related adverse events. Overall, 70% of patients reported at least one adverse event. This rate is typical in a cohort of critically ill patients. Only 11 adverse events were related to the CP transfusion, and four of these were identified as SAEs. All 11 adverse events were classified as “possibly related” to the intervention by the treating physician using a four-item Likert-type scale (probably, possibly, remotely, or not related). TRALI was the most frequent SAE (1.5%, n = 3). This incidence is within the 0.001 to 2.1% range reported in both the general population and critically ill patients (**34, 35**).

In patients with COVID-19 especially, it is difficult to assign with certainty the occurrence of TRALI or Transfusion-associated circulatory overload (TACO) since, in the context of mostly severe cases, respiratory failure is the predominant dysfunction and concomitant but potentially unrelated worsening may occur close to the transfusion. As for thrombocytopenia, post-transfusion purpura is rare, with an incidence of less than 0.01%, associated with the detection of antiplatelet antibodies (**36**). In the only case of thrombocytopenia, the antiplatelet antibody test was negative. Different mechanisms likely underlie COVID-19-related thrombocytopenia, including direct inhibition of hematopoiesis in bone marrow by SARS-CoV-2, autoimmune destruction induced by the virus, and platelet aggregation and consumption in the lung parenchyma (**37**).

The present study has some limitations. Since we aimed to assess mortality rates and safety, the absence of a control group prevents drawing conclusions on the benefits of CP treatment. The lack of a control group was grounded on ethical concerns raised at the beginning of the outbreak when the study was designed. Nevertheless, the observed mortality rates are encouraging and additional studies with matched control cases are planned. While donor anti-SARS-CoV-2 spike protein antibody titers were elevated (≥ 1:320), we have not yet investigated their neutralizing activity. Ongoing experiments aim to characterize the neutralizing antibody and cytokine content of CP and identify the factors potentially responsible for its therapeutic benefits in patients with COVID-19 using multivariate analysis. Such studies will generate new hypotheses on CP efficacy in regards to specific types and amounts of neutralizing antibodies and cytokines.

Although CP transfusion has shown promising results against COVID-19 in case series (18,19) and case-control studies (**38**), its real benefit remains unclear. The first published randomized trial with CP in COVID-19 did not show significant improvement in the clinical condition of patients with severe, life-threatening COVID-19 but the interpretation is limited by the study’s early endpoint. Nevertheless, patients with severe, non-life-threatening COVID-19 improved significantly compared with the control group (91.3% vs. 68.2% (HR, 2.15 [95% CI, 1.07-4.32]; p = 0.03) (**39**). These findings suggest that CP may be more effective in patients with COVID-19 without life-threatening conditions.

## Conclusions

CP is safe when used in the COVID-19 population even for those who present severity criteria and/or risk factors for poor prognosis including cancer. The mortality rate reported here is comparable to those found in larger series and was higher in patients requiring ventilatory support and those over 65 years old. In-depth analyses of the serological and molecular characteristics of CP are needed to evaluate the efficacy of this intervention through controlled clinical trials.

## Data Availability

We are unable to make the data available in a public repository or uploaded as supplementary information because this is not permitted by our organisation s research governance policy and ethics committee regulations. Anonymised data can be made available to researchers who meet the conditions of the ethics approval and research governance policy that applies to this study. Researchers may request anonymized data access by contacting Principal Investigator at

## Acknowledgments

All study coordinators in particular to Giselle Godoy, Cecilia Martinez, Macarena Ubal, Mayra Doddis, Isabel Rojo, and Ricardo Palomera. Blood Bank staff from participating hospitals: Marcelo Díaz de Valdes, Jaime Pereira, Mayling Chang, Pablo Sepulveda, Ricardo Hojas, Veronica Soto, Eugenio Reyes, Ana Gonzalez, Celia Gamonal, Marcos Espinoza, Marcos Sandoval, and Marcelo Vega. Nordiana Baruzzi and Luz María González Anguita. We’d like to thank the catg.cl laboratory for sampling processing at Punta Arenas, XII Region. Chile. M.N. is supported by Grant MAG 1895. M.E.B is supported by Fondo Nacional de Desarrollo Científico y Tecnológico.

